# Gut microbiome and incidence of major adverse cardiovascular events in the general population: a prospective study in five European cohorts

**DOI:** 10.64898/2026.11.10.26367787

**Authors:** Oleg Kambur, Louise Grahnemo, Maria Nethander, Kamalita Pertiwi, Koen F. Dekkers, J. Gustav Smith, Gunnar Engström, Johan Ärnlöv, Marju Orho-Melander, Robert F.J. Kullberg, Bastaan W. Haak, Henrike Galenkamp, Bert-Jan van den Born, Max Nieuwdorp, Kari Koponen, Guillaume Méric, Rob Knight, Teemu Niiranen, Kristian Hveem, Tove Fall, Susanna C. Larsson, Claes Ohlsson, Veikko Salomaa, Aki S. Havulinna

**Author notes:** Corresponding author,. Address: Department of Medicine, University of Eastern Finland. Yliopistonrinne 3 (P. O. Box 1627). 70211 KUOPIO.

## Abstract

**Background:** Accumulating evidence suggests that the gut microbiota is implicated in cardiovascular disease. We investigated how the gut microbiome associates with incident major adverse cardiovascular events (MACE), and cardiovascular and all-cause mortality (ACM) in a large, prospective, population-based study. The findings were tested for replication in four independent cohorts.

**Methods:** Gut microbiome composition was assessed via shallow shotgun sequencing in the FINRISK 2002 cohort, with up to 18 years of follow-up. Associations of microbial taxa with incident MACE were investigated using time-to-event analyses. Significant associated microbial features were tested for replication in four European cohorts (HUNT, HELIUS, SIMPLER, and SCAPIS) followed by a meta-analysis.

**Results:** In the FINRISK 2002 cohort (1167 incident MACE events), nine taxa were significantly associated with incident MACE and sustained adjustments for traditional risk factors and multiple comparisons. The meta-analysis of replication cohorts (N=30,228; 1,938 participants with incident MACE) confirmed robust associations for several identified taxa. Specifically, higher abundance of genera *Scardovia* (HR=1.17 [95% CI 1.08-1.26]*, p*= 0.0001) and *Limosilactobacillus* (HR=1.16 [95%CI 1.09-1.23]*, p*= 1E-06) was associated with increased MACE risk, and increased risk of cardiovascular mortality and all-cause mortality. The species *ER4 sp900317525* (HR=0.89 [95%CI 0.83-0.95], *p*=0.0005) was associated with lower hazard of MACE and all-cause mortality.

**Conclusions:** The gut metagenome showed robust associations with incident MACE, which were replicated in four independent European cohorts. Key microbial taxa also associated with incident MACE and mortality, suggesting their potential relevance as candidate risk markers for further studies.

**Clinical Perspective:** *What is New?:*

- In a large-scale discovery and replication study involving five European population-based cohorts (> 36,000 participants) we identified specific gut microbial taxa—including the genera *Scardovia* and *Limosilactobacillus* - which were significantly associated with increased risk of major adverse cardiovascular events (MACE).
- The obligate anaerobic commensal *ER4 sp900317525*, member of the *Oscillospiraceae* family, was associated with a lower risk of incident MACE across independent cohorts.
- These microbial signatures remained significant after adjusting for traditional cardiovascular risk factors and were further associated with both cardiovascular and all-cause mortality.

*What Are the Clinical Implications?:*

- The gut microbiome may represent a novel and modifiable risk factor for MACE as well as cardiovascular- and all-cause mortality.
- These findings provide a rationale for future clinical trials investigating whether targeted modulation of the gut microbiome—through diet, prebiotics, oral hygiene, or other targeted therapies—can reduce the risk of MACE and improve patient survival.

## Introduction

Cardiovascular disease (CVD) remains the leading cause of morbidity and mortality worldwide, accounting for nearly 18 million deaths annually and imposing a substantial burden on global health and healthcare systems.^1^ While established risk factors, such as hypertension, dyslipidemia, diabetes, smoking, and obesity, explain a large proportion of cardiovascular risk, they identify only about half of individuals who subsequently experience major adverse cardiovascular events (MACE).^2^ Persistent residual risk, even among patients receiving guideline-directed therapy, underscores the need to identify novel biological pathways and biomarkers that contribute to cardiovascular event susceptibility.^3^

The gut microbiome has emerged as a critical modulator of host processes central to atherosclerosis, thrombosis, and myocardial injury – metabolism, immune function, and systemic inflammation.^4,5^ Comprising trillions of microorganisms and their genes, the gut microbiome produces a wide range of bioactive metabolites such as short-chain fatty acids (SCFAs), secondary bile acids, trimethylamine-derived metabolites, and tryptophan catabolites. These metabolites co-influence lipid metabolism, endothelial function, vascular tone, platelet reactivity, and immune homeostasis.^6,7^

Disruption of this ecosystem, dysbiosis, is characterized by reduced microbial diversity, altered taxonomic composition, and impaired functional capacity, and has been associated with hypertension, atherosclerosis, coronary artery disease, atrial fibrillation, myocardial infarction, stroke, and heart failure, as shown by sequencing-based studies.^8–12^ Together, these studies suggest that gut microbiome composition may contribute to cardiovascular events.

However, because patients with CVD frequently have multiple risk factors that are themselves associated with gut microbiota alterations, the cross-sectional design of these studies makes it impossible to disentangle microbiota characteristics predictive of MACE from changes related to underlying risk factors, and therefore to determine whether microbiota alterations precede and contribute to CVD development or merely arise as a consequence of the acute event and their treatments. Moreover, current studies are characterized by substantial heterogeneity in analytical approaches and often lack independent replication. As a result, the clinical relevance and reproducibility of microbiome–cardiovascular disease associations have remained uncertain.

To address this, we aimed to identify gut microbiota signatures (present before the event) associated with the risk of incident MACE in the general population, using a well-characterized cohort with longitudinal follow-up. We subsequently validated these signatures in four independent population-cohorts including over 30,000 participants. By integrating microbiome profiling with time-to-event analyses for future cardiovascular events, this study aims to clarify the role of the gut microbiome in cardiovascular event risk, and to inform of its potential utility in future preventive strategies.

## Materials and methods

### Study design

We conducted this study in five independent population-based cohorts that combined gut microbiota characterization with long-term follow-up on the incidence of MACE following fecal sample collection. The FINRISK 2002 cohort was used as discovery cohort. The Trøndelag Health Study (HUNT, Helseundersøkelsen i Nord-Trøndelag, Norway), the Healthy Life in an Urban Setting cohort (HELIUS, the Netherlands), the Swedish Infrastructure for Medical Population-Based Life-Course and Environmental Research cohort (SIMPLER) and the Swedish CArdioPulmonary bioImage Study (SCAPIS) served as validation cohorts.

### Cohort descriptions

#### Discovery cohort - FINRISK 2002

##### General

FINRISK 2002 is a health examination survey based on a random and representative population sample. Details on study design, in- and exclusion criteria and follow-up have been previously described^13,14^ and are provided in the Supplementary Material. In summary, participants were sampled from the National Population Information System and stratified by sex, region and 10-year age groups. Participants responded to questionnaires, underwent physical examination by trained nurses, and gave blood samples. Participants with complete clinical and microbiome data who permitted registry data access were included in the study (**Fig.1**). Participants with prevalent MACE at baseline (n=370) were excluded. The final sample thus included 6,632 persons, of whom 5,471 also had genotype data. The FINRISK 2002 study was approved by the Coordinating Ethics Committee of the Helsinki and Uusimaa University Hospital District (#558/E3/2001), and all participants gave written informed consent.

**Figure 1.**
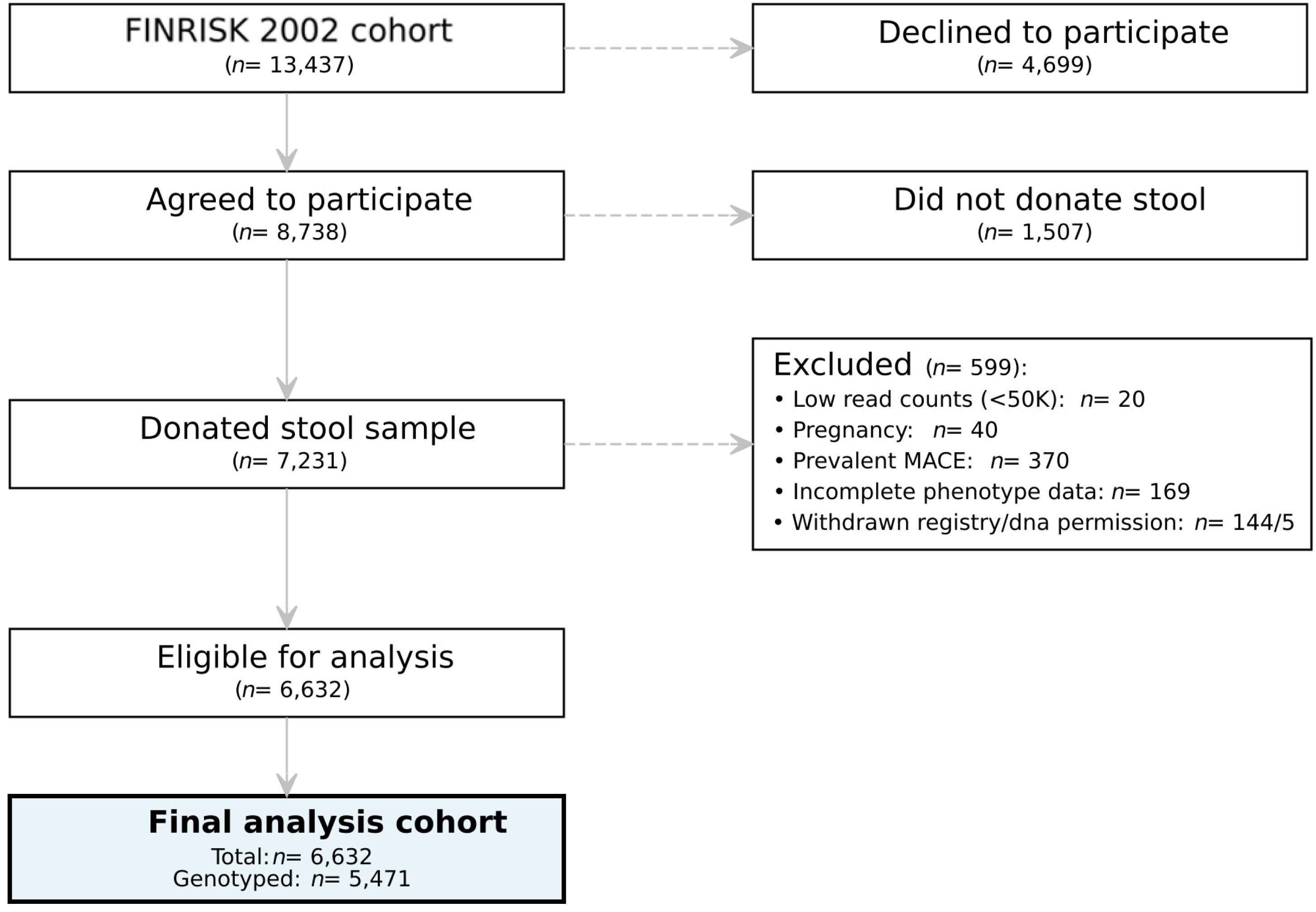
Flowchart of the discovery cohort.

##### Outcomes and cardiovascular endpoints

Our main outcomes were incident (first ever) MACE (Major Adverse Cardiovascular Event) as a primary outcome and cardiovascular mortality and all-cause mortality as secondary outcomes. MACE was defined as non-fatal or fatal CHD event, coronary revascularization, non-fatal or fatal ischemic stroke and hospitalized or fatal heart failure, whichever happened first. These events were identified from the care register for health care, heart failure was additionally identified from the drug purchase register and deaths from the causes of death register. These registers ensure practically complete coverage of all major health events, including cardiovascular events, and deaths, over an individual’s lifetime. The reliability, validity and completeness of the Finnish health care registers have been extensively documented.^15–17^ The International Classification of Diseases, 9 and 10 codes used for MACE are presented in **supplementary table S1**, along with Nordic Medico-Statistical Codes (NOMESCO) used for identifying revascularizations. The participants of the current study have been followed through December 31, 2019.

#### Replication cohorts

Replication cohorts included HUNT, HELIUS, SIMPLER and SCAPIS. Descriptions of the replication cohorts, their MACE definitions, applied exclusion and inclusion criteria, sample sizes, statistical methods and metagenome descriptives are given in the supplements (**S2: Other supplementary methods, S1: MACE definitions, S3: Replication cohort descriptives, S4: Metagenome descriptives, S5: Sample sizes)**. Importantly, the definition of MACE, applied exclusion and inclusion criteria, and statistical methods for the replication cohorts closely followed those of FINRISK.

## Materials and methods overview

Details on used clinical variables, fecal sample collection, DNA extraction, microbiota sequencing, and preprocessing as well as genetic data and polygenic risk score (PRS) protocols have been previously described and are provided in the supplement (**S2: Other supplementary methods)**.^13,18–24^ Metagenome descriptives for FINRISK are shown in supplementary (**Table S6**).

### Statistical analysis

All statistical analyses were performed using R (version 3.6.3 and 4.5.2). In statistical tests, a nominal two-tailed p<0.05 was considered statistically significant. To account for multiple testing, p-values were adjusted using Benjamini-Hochberg false discovery rate (FDR) procedure, as indicated below. Significance threshold was set at FDR (Q) < 0.05.

We prespecified three key features of the bacterial gut microbiome to assess their associations with the risk of MACE: 1) alpha diversity (Shannon index at species-level count data), 2) overall gut microbiota composition (beta-diversity with Aitchison distances calculated as Euclidian distances of the central log ratio (CLR) transformed relative abundances followed by principal component analysis on the Euclidean Aitchison distance matrix), , and 3) individual taxa associations. Individual taxa (CLR-transformed relative abundances) were analyzed on three taxonomic levels: phylum, genus and species. To eliminate mathematical singularities from zero values in CLR-transformation, a global pseudocount equal to half of the minimum observed non-zero value across the entire dataset was used. No prevalence or abundance filtering was used.

Cox proportional hazards regression was used to assess the association of microbial features with incident MACE. All models were stratified by sex, and age was used as the time-scale. No transformations were used for the covariates, unless otherwise noted. Individuals were censored at death due to other causes, except in the all-cause mortality analyses, or at the full observation time (18 years for the FINRISK discovery cohort). Cox model assumptions were checked using the *cox.zph* function in R. After considering corrections for multiple testing, all model assumptions were met. Effect sizes are expressed *per* standard deviation.

Several **measures of microbial composition** were tested for association with incident MACE, including alpha-diversity, principal components of beta-diversity, as well as individual taxa.

We used three Cox regression models with increasing levels of adjustment. A minimal *demographically-adjusted* model (model 1) using age as the time scale and stratifying for sex was used as the reference model. Prevalent use of medication with potential confounding impact on the microbiome was used as a binary covariate. Multiple comparison FDR corrections were applied to the model 1 results adjusting for the number of analyzed taxa (N = 4839, 1497 and 33 for species, genera, and phyla, respectively).

To further assess the potential impact of traditional cardiovascular risk factors, the analyses were repeated using the *clinically-adjusted model (model 2*). This model also included BMI, smoking status, systolic blood pressure, total and HDL cholesterol, antihypertensive medication and prevalent diabetes as covariates.

To assess the potential confounding impact of genetic risk for coronary artery disease, the analyses were repeated using the PRS-adjusted model (model 3). This model included the same traditional cardiovascular risk factors as model 2, and the CAD PRS, 10 principal components of the genotype data and genotyping batch. PRS and PCs were standardized to have mean=0, SD=1.

For replication we considered a nominal p-value < 0.05 with concordant directionality as supportive evidence. A nominal p-value < 0.05 with concordant directionality in meta-analysis was considered as formal replication.

### Meta-analysis

We performed meta-analysis of replication cohorts. Minimally-adjusted model was considered (see **S2: Other supplementary methods** for details). We combined the cohort-specific hazard ratios (HRs) using both fixed-effect (inverse variance weighting) and random-effects meta-analysis (restricted maximum likelihood, REML). We derived log hazard ratios (logHR) and standard errors (SE) from Cox regression results for each cohort. We report the results of fixed-effects meta-analysis unless significant heterogeneity was observed (*I²* > 50%, *Q-*test *p-*value*, Q_p_ <*0.1). Finally, Meta-analysis of all cohorts was performed to obtain pooled effect size estimates.

### Sensitivity analyses: sex and proton pump inhibitors

In sensitivity analyses we assessed the effects of proton pump inhibitors (PPI) and sex in FINRISK. Effects of PPI were assessed by using PPI-adjusted minimal model. Effects of sex were assessed by sex-specific analyses where males and females were analysed separately.

### Secondary analyses: cardiovascular and all-cause mortality

For taxa showing significant associations with MACE in FINRISK and meta-analysis, we further evaluated their associations with cardiovascular and all-cause mortality.

Mortality analyses were performed in the full study sample, including participants with prevalent MACE. Cox proportional hazards models were fitted as in the main analyses, using model 1.

## Results

### Characteristics

The discovery cohort comprised 6,667 participants, including 1,167 individuals with incident MACE and 5,500 individuals with no MACE; 371 individuals with prevalent MACE were excluded. The sample included 3,704 females and 2,963 males (**Table 1**).

**Table 1.**
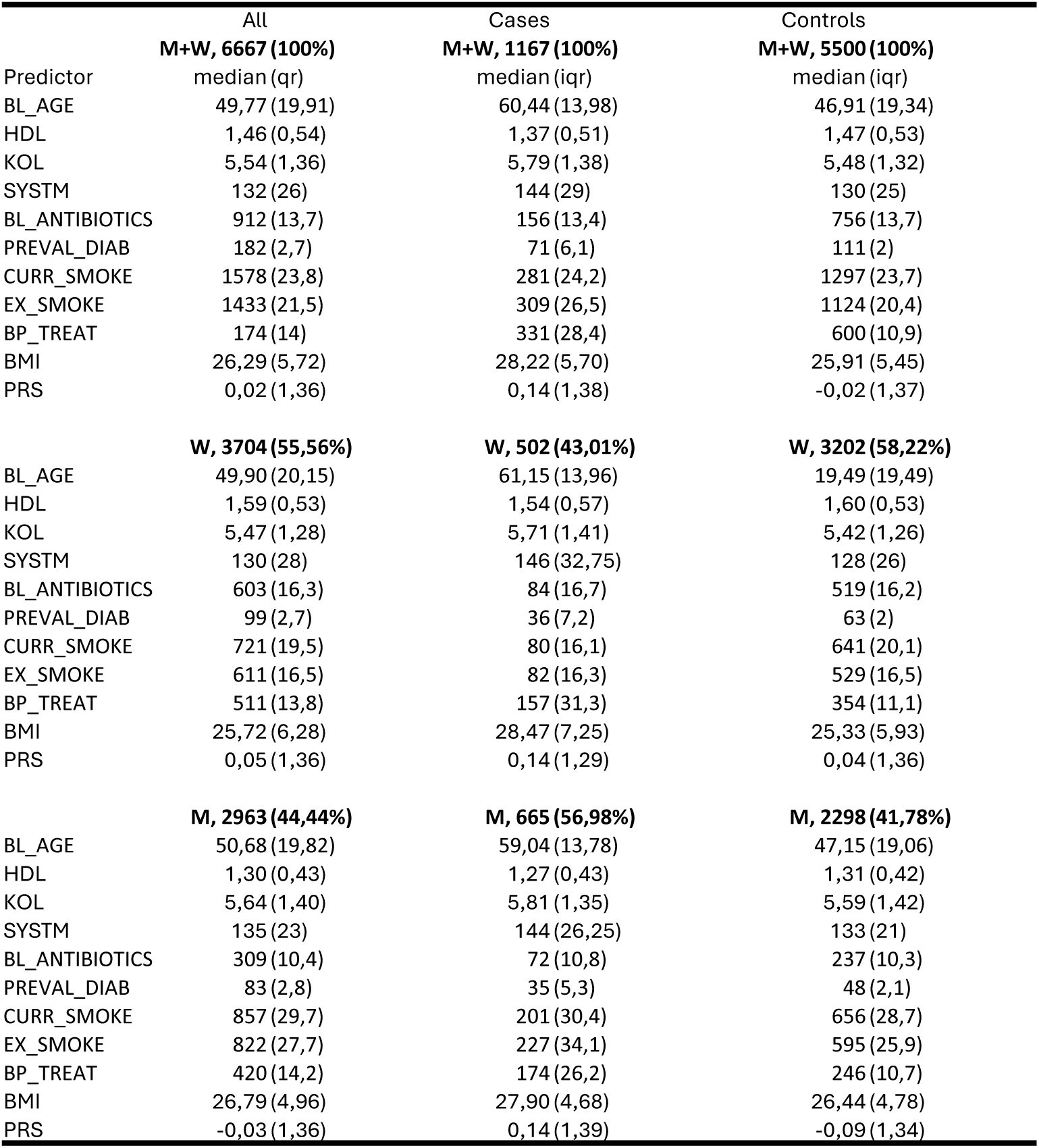
FINRISK cohort descriptives. Numbers are calculated from the phenotype data prior to the exclusions. In the final analyses there were 6632 participants with complete data for minimal model. PRS data was missing from 16% of participants. For discrete variables n(%) was calculated by category totals. BL_AGE, age at baseline; HDL, high-density lipoprotein cholesterol; KOL, total cholesterol; SYSTM, systolic blood pressure; BL_ANTIBIOTICS, antibiotic use within 6 months before baseline; PREVAL_DIAB, prevalent diabetes; CURR_SMOKE, current smoker; EX_SMOKE, ever smoker; BP_TREAT, blood pressure medication use; BMI, body mass index; PRS, polygenic risk score for coronary artery disease.

|  | All | Cases | Controls |
|---|---|---|---|
|  | **M+W, 6667 (100%)** | **M+W, 1167 (100%)** | **M+W, 5500 (100%)** |
| Predictor | median (qr) | median (iqr) | median (iqr) |
| BL_AGE | 49,77 (19,91) | 60,44 (13,98) | 46,91 (19,34) |
| HDL | 1,46 (0,54) | 1,37 (0,51) | 1,47 (0,53) |
| KOL | 5,54 (1,36) | 5,79 (1,38) | 5,48 (1,32) |
| SYSTM | 132 (26) | 144 (29) | 130 (25) |
| BL_ANTIBIOTICS | 912 (13,7) | 156 (13,4) | 756 (13,7) |
| PREVAL_DIAB | 182 (2,7) | 71 (6,1) | 111 (2) |
| CURR_SMOKE | 1578 (23,8) | 281 (24,2) | 1297 (23,7) |
| EX_SMOKE | 1433 (21,5) | 309 (26,5) | 1124 (20,4) |
| BP_TREAT | 174 (14) | 331 (28,4) | 600 (10,9) |
| BMI | 26,29 (5,72) | 28,22 (5,70) | 25,91 (5,45) |
| PRS | 0,02 (1,36) | 0,14 (1,38) | -0,02 (1,37) |
|  | **W, 3704 (55,56%)** | **W, 502 (43,01%)** | **W, 3202 (58,22%)** |
| BL_AGE | 49,90 (20,15) | 61,15 (13,96) | 19,49 (19,49) |
| HDL | 1,59 (0,53) | 1,54 (0,57) | 1,60 (0,53) |
| KOL | 5,47 (1,28) | 5,71 (1,41) | 5,42 (1,26) |
| SYSTM | 130 (28) | 146 (32,75) | 128 (26) |
| BL_ANTIBIOTICS | 603 (16,3) | 84 (16,7) | 519 (16,2) |
| PREVAL_DIAB | 99 (2,7) | 36 (7,2) | 63 (2) |
| CURR_SMOKE | 721 (19,5) | 80 (16,1) | 641 (20,1) |
| EX_SMOKE | 611 (16,5) | 82 (16,3) | 529 (16,5) |
| BP_TREAT | 511 (13,8) | 157 (31,3) | 354 (11,1) |
| BMI | 25,72 (6,28) | 28,47 (7,25) | 25,33 (5,93) |
| PRS | 0,05 (1,36) | 0,14 (1,29) | 0,04 (1,36) |
|  | **M, 2963 (44,44%)** | **M, 665 (56,98%)** | **M, 2298 (41,78%)** |
| BL_AGE | 50,68 (19,82) | 59,04 (13,78) | 47,15 (19,06) |
| HDL | 1,30 (0,43) | 1,27 (0,43) | 1,31 (0,42) |
| KOL | 5,64 (1,40) | 5,81 (1,35) | 5,59 (1,42) |
| SYSTM | 135 (23) | 144 (26,25) | 133 (21) |
| BL_ANTIBIOTICS | 309 (10,4) | 72 (10,8) | 237 (10,3) |
| PREVAL_DIAB | 83 (2,8) | 35 (5,3) | 48 (2,1) |
| CURR_SMOKE | 857 (29,7) | 201 (30,4) | 656 (28,7) |
| EX_SMOKE | 822 (27,7) | 227 (34,1) | 595 (25,9) |
| BP_TREAT | 420 (14,2) | 174 (26,2) | 246 (10,7) |
| BMI | 26,79 (4,96) | 27,90 (4,68) | 26,44 (4,78) |
| PRS | -0,03 (1,36) | 0,14 (1,39) | -0,09 (1,34) |

Baseline characteristics are summarized in **Table 1**. In brief, individuals who developed MACE had higher systolic blood pressure, total cholesterol, BMI, lower HDL cholesterol and higher PRS compared with MACE-free participants. Diabetes and smoking were more prevalent among individuals who developed MACE.

### Replication cohorts

In the replication cohorts, a total of 30,228 participants were included of whom 1,938 developed MACE during follow-up and 28,290 did not. Specifically, in **HUNT** there were 11,604 persons in total, including 635 individuals who developed MACE and 10,969 individuals who did not. In **HELIUS** there were 4,248 persons in total, including 126 individuals who developed MACE and 4,122 individuals who did not. In **SIMPLER,** 5,192 persons were free of MACE at baseline, among whom 881 developed incident MACE during follow-up. In **SCAPIS** there were 9,184 persons in total, of whom 296 individuals developed incident MACE. Altogether our study included 2,805 individuals who experienced incident MACE and 34,055 individuals who did not. Replication cohorts are described in more detail in the supplement (**Supplementary table S3, Supplementary methods S2)**.

## MAIN RESULTS

### Overall gut microbiota composition and risk of MACE

Shannon **alpha diversity** was not associated with incident MACE in any of the models (*p* > 0.05 for all comparisons, **Table S7**). Notably, the lack of association was annotation-specific and sensitivity analyses using *gtdb*-annotation revealed statistically significant inverse association between alpha diversity and MACE risk.

#### Beta-diversity

The first principal component (PC) explained over 75% of variance in Aitchison beta diversity. The cumulative proportion of explained variance for the first ten PCs reached 90%. Incident MACE was associated with Aitchison PCs 6 and 7 (**Table S7**). The association with Aitchison PC6 sustained adjustments for <u>traditional</u> <u>cardiovascular risk factors</u> and remained significant in the clinically- and genotype-adjusted models (**Table S7**).

### Associations of individual taxa in FINRISK

#### MACE

In initial analysis of SHOGUN-annotated individual taxa, we found five bacterial taxa that were significantly associated with an increased risk of MACE during follow-up, after multiple comparison corrections (FDR) (**Fig. 2, Table S8, S9**): *Scardovia* at the Genus level, and the species *Scardovia wiggsiae*, *Lactobacillus gasseri*, *Lactobacillus gastricus*, and *Lactobacillus antri*. In addition to age, sex and prior use of medication with impact on gut microbiota, these associations also sustained correction for traditional cardiovascular risk-factors (BMI, smoking status, systolic blood pressure, total and HDL cholesterol, antihypertensive medication and prevalent diabetes) in a multivariable model. Moreover, all except *L. antri* remained robust after adjusting for the polygenic risk score (PRS) (**Table S8**).

**Figure 2.**
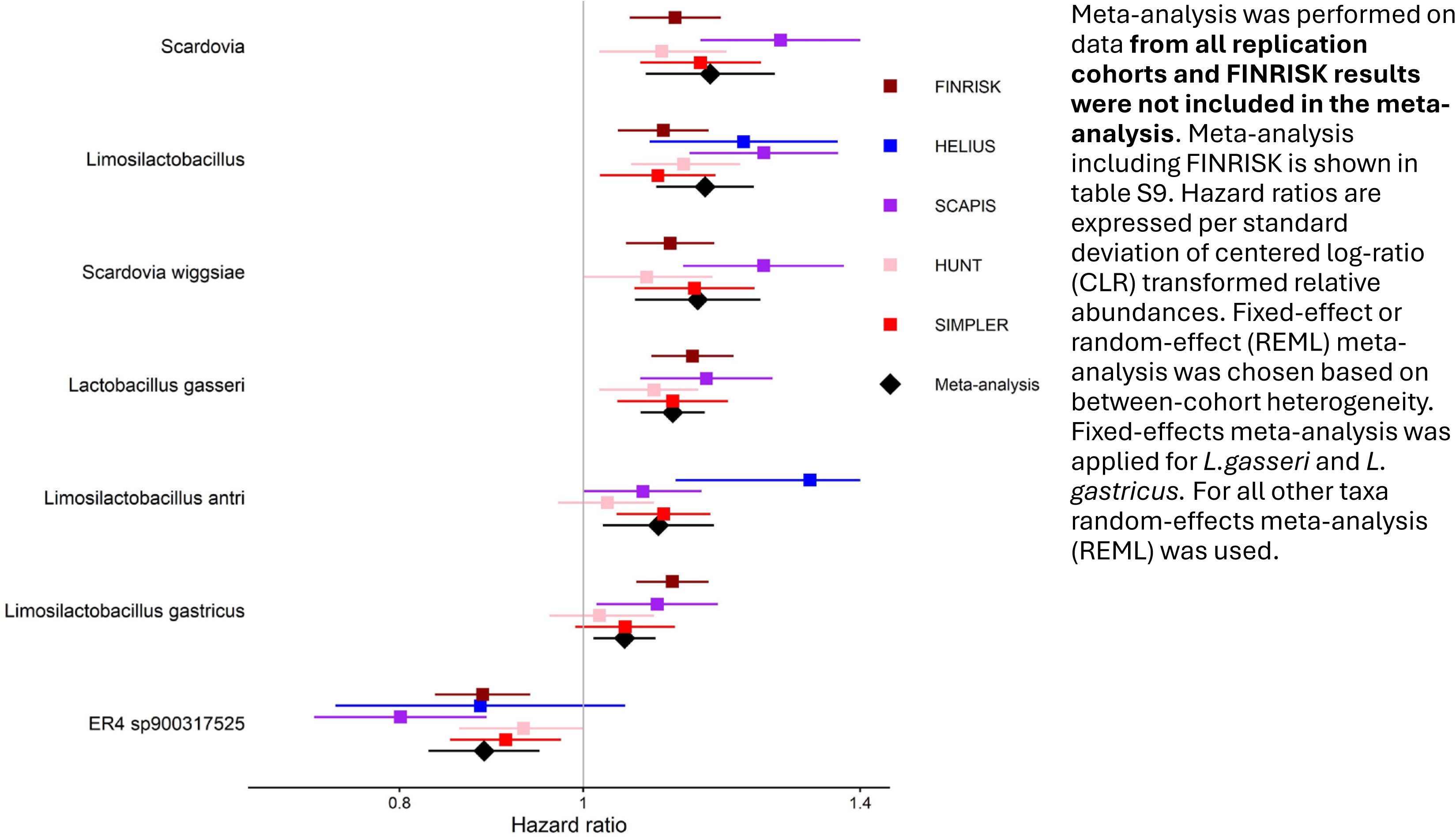
Prospective associations of taxa with incident MACE: FINRISK results, replication and meta-analysis. Meta-analysis was performed on data from all replication cohorts and FINRISK results were not included in the meta-analysis. Meta-analysis including FINRISK is shown in table S9. Hazard ratios are expressed per standard deviation of centered log-ratio (CLR) transformed relative abundances. Fixed-effect or random-effect (REML) meta-analysis was chosen based on between-cohort heterogeneity. Fixed-effects meta-analysis was applied for L.gasseri and L. gastricus. For all other taxa random-effects meta-analysis (REML) was used.

Analyses of Greengenes2-annotated microbiome revealed three additional taxa associations, including *ER4 sp900317525* (HR = 0.88 [95%CI: 0.84-0.94], *p=*3.46E-05, *Lachnospira eligens A* (HR = 0.87 [95%CI: 0.82-0,92]), *p* =7.26E-07 and *Lachnospira* (HR = 0.89 [95%CI: 0.84-0.94], p = 1.83E-05).

### Replication and meta-analysis

Individual taxa associated with MACE in FINRISK were evaluated in four independent cohorts. Despite differences in taxa prevalences between cohorts (**TableS4, S6**), 7 associations were replicated with statistically significant results (**Fig. 2, Table S9**). For associations which did not reach significance, the effect estimates were directionally consistent with the FINRISK findings, with the exception of *Lachnospira* in HUNT.

All replication cohorts were included in the meta-analysis, producing altogether 1,938 incident MACE events (**Table S9**). Heterogeneity across different taxa results ranged from negligible to substantial (*I^2^* = 0-81%, *Q_p_*=0.46 to <0.01, tau^2^=0-0.003).

In meta-analysis on **genus** level, including all replication cohorts, both *Limosilactobacillus* and *Scardovia,* were associated with increased risk of MACE (*Limosilactobacillus:* HR=1.16 [95%CI 1.09-1.23]*, p*=1.00E-06; *Scardovia:* HR=1.17 [95% CI 1.08-1.26]*, p*= 0.0001; **Table S9**). *Lachnospira* was not significant.

On the **species** level, *S. wiggsiae, L. gasseri, L. gastricus* and *L. antri* were associated with incident MACE (HR=1.04-1.15 [95%CI: 1.01-1.24], *p=-*5.06E-088.98E-03; **Fig. 2, Table S9**). *Lachnospira* and *Lachnospira eligens A* showed similar directional trends to the FINRISK cohort, but lacked statistical significance. *ER4 sp900317525* showed inverse association (HR=0.89 [95%CI 0.83-0.95], *p*=4.76E-04; **Fig. 2, Table S9**). In pooled meta-analysis of all cohorts including FINRISK estimates were in line with estimates of meta-analysis of replication cohorts.

### Secondary analyses: mortality

Cardiovascular and all-cause mortality were assessed in the FINRISK 2002 cohort.

#### Cardiovascular mortality

*Scardovia* and *Limosilactobacillus* were associated with increased risk of cardiovascular mortality (HR=1.08 [95%CI:1.03–1.13], *p=*0.0011 and HR=1.08 [95%CI:1.03-1.14], *p=*0.0024 respectively; **Fig. 3, Table S10**).

**Figure 3.**
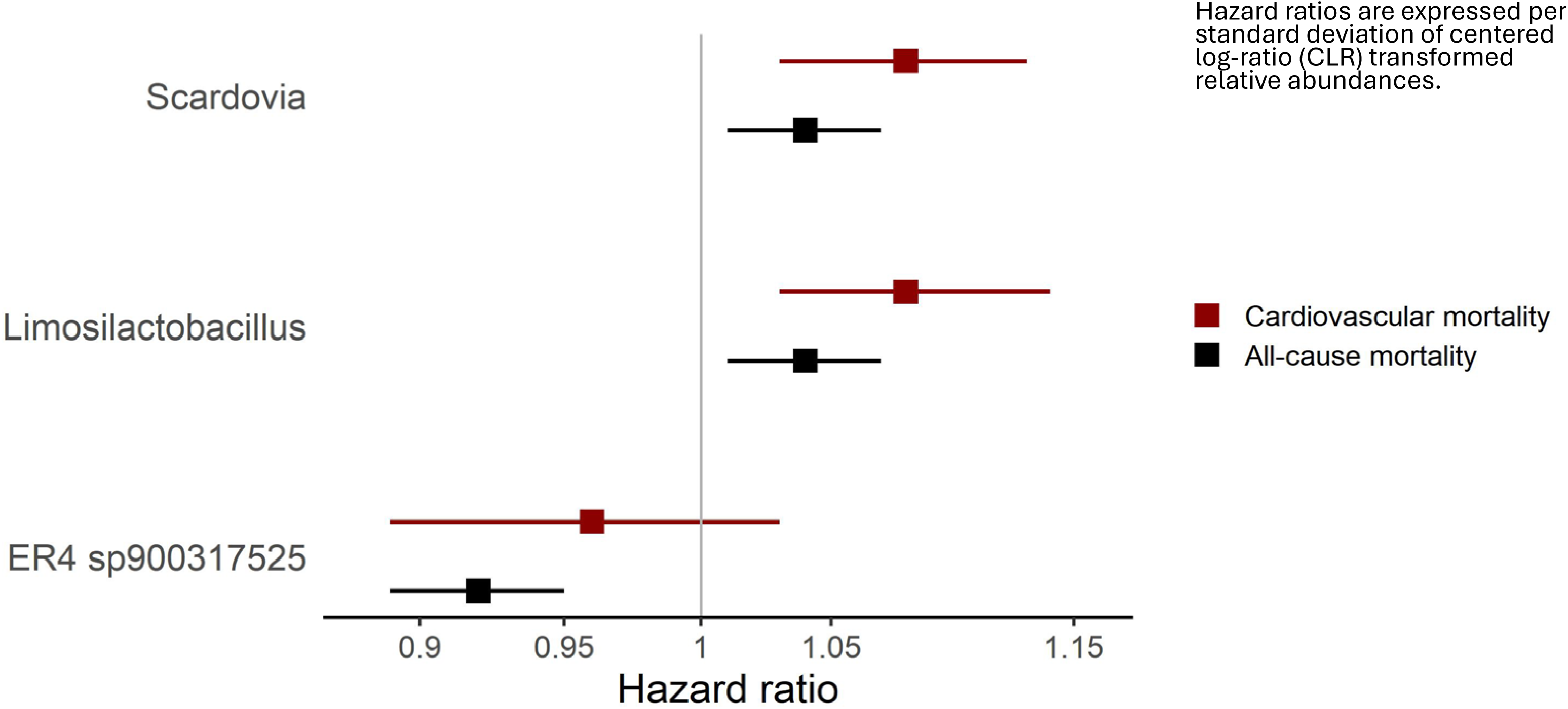
Associations of taxa with cardiovascular and all-cause mortality in FINRISK. Hazard ratios are expressed per standard deviation of centered log-ratio (CLR) transformed relative abundances.

#### All-cause mortality

*Scardovia* genus was associated with increased risk of all-cause mortality (HR=1.04 [95%CI:1.01–1.07], *p=*0.0028; **Fig. 3, Table S10).** *Limosilactobacillus* showed similar results (HR=1.04 [95%CI:1.01–1.07], *p=*0.0069; **Fig. 3, Table S10**).

#### ER4 sp900317525

*ER4 sp900317525* was associated with lower all-cause mortality (HR=0.92 [95% CI: 0.89–0.95], p=2 × 10⁻⁶; 861 events), but not with cardiovascular mortality (HR=0.96 [95% CI: 0.89–1.03], *p*=0.21; 241 events; **Fig.3**).

As *ER4 sp900317525* was not associated with cardiovascular mortality, we further examined non-cardiovascular causes of death. *ER4 sp900317525* was associated with **lower cancer mortality** (HR=0.93 [95% CI: 0.87–0.99], *p*=0.025 237 events) and with **lower mortality from other non-cardiovascular causes** (HR=0.91 [95% CI: 0.85– 0.98], p=0.0089; 213 events).

### Sensitivity analyses

In sensitivity analyses that adjusted for PPI usage, all associations remained significant **(Table S11)**. In analyses with sex interaction term, sex interaction was not significant for any of the taxa. When males and females were analysed separately, all associations remained directionally similar and statistically significant **(Table S11)**, with the exception for *ER4_sp900317525* in males (HR=0.93 [CI95% 0.86-1.01], p=0.068) and *Limosilactobacillus* in females (HR=1.08, [CI95% 0.99-1.18], p=0.086) which no longer reached statistical significance although the effect estimates remained consistent with those from the main analysis. Interestingly, *ER4_sp900317525* showed a trend toward sexual dimorphism and stronger effect in females (HR=0.84 [CI95% 0.77-0.91], p=3.06e-05).

Similar effect sizes, albeit with lower precision, were observed in additional SCAPIS sensitivity analyses after excluding participants with recent antibiotic use and adjusting for additional SCAPIS-specific covariates (**Table S12**).

## Discussion

### Main findings and added value of the study

In our prospective study of altogether 36,860 individuals and 2,805 cases of incident MACE, we identified 9 microbial taxa associated with incident MACE. For all 9 taxa the existing prior evidence was scarce. The results remained robust after adjusting for traditional cardiovascular risk factors and CHD PRS. This independence suggests that the observed association is not merely a byproduct of established clinical risks or genetic predisposition, but rather represents a distinct, independent risk factor.

For 7 of the 9 taxa our findings were successfully replicated in meta-analysis of four replication cohorts. Notably, the microbial taxa also showed consistent associations with cardiovascular and all-cause mortality, underscoring potential clinical importance of these microbial signatures in long-term cardiovascular outcomes.

Clinically, these findings highlight a potential for risk assessment, suggesting that evaluating microbiome could provide unique insights alongside standard cardiovascular workups. Because the gut microbiome is modifiable through diet, lifestyle, or targeted therapeutics, these findings highlight a potential novel avenue for reducing MACE risk. However, future interventional studies are required to establish causality and evaluate whether modulating gut microbiome can directly improve clinical outcomes.

### Scardovia

*Scardovia wiggsiae* (SW) which belongs to *Scardovia* genus is a relatively recently identified gram-positive, anaerobic bacterium that has emerged as a significant **oral pathogen**. It has been linked to severe dental caries,^25^ and gingivitis. *Scardovia* is acidogenic and acid-tolerant and usually occurs on surfaces of teeth, tongue and gingival crevices as well as in saliva. Both *Scardovia* genus and *S. wiggsiae* species were associated with increased risk of incident MACE.

### Lactobacillus and Limosilactobacillus

Several *Limosilactobacillus* species (*L. gasseri*, *L. gastricus*, *L. antri*) were robustly associated with incident MACE. These taxa were previously classified within the *Lactobacillus* genus. Recently, *Lactobacillus* genus was reorganized and *L. gastricus* and *L. antri* reassigned to *Limosilactobacillus*, reflecting updated phylogenetic resolution.^26^

At the broader *Lactobacillus* genus level, multiple species showed nominal associations with MACE which did not withstand correction for multiple comparisons. *Lactobacilli* were predominantly associated with increased MACE risk, except for *L. thailandensis* and *L. rhamnosus*, a commonly used probiotic species, which were nominally associated with reduced MACE risk. Additionally, several species (*L. vaginalis*, *L. nagelii*, *L. lindneri*) were linked to beta-diversity components associated with increased MACE risk, further suggesting heterogeneous, species-specific effects within the genus. This is in line with our earlier study showing heterogeneity among *Lactobacillus* species in blood pressure associations.^27^ This heterogeneity likely explains the absence of significant associations at the *Lactobacillus* genus level. In contrast, the association observed for the *Limosilactobacillus* genus suggests a more functionally or ecologically coherent group of species contributing to cardiovascular risk.

Collectively, these findings suggest that rather than being benign bystanders, elevated *Lactobacilli* and *Limosilactobacilli* may function as active players in systemic inflammatory processes contributing to MACE. This view is further supported by association of increased *Lactobacillus* abundances in saliva with increased overall mortality in an earlier study^28^ and our results showing association of increased abundances of *Limosilactobacillus* in the gut and all-cause mortality.

### ER4 sp900317525

The obligate anaerobic commensal *ER4 sp900317525* is a member of the *Oscillospiraceae* family of *Clostridia* class. Our study associates *ER4 sp900317525* with lower risk of cardiovascular events (MACE). Earlier evidence for association of *ER4 sp900317525* with **cardiovascular health** is limited. Previous research have suggested that *Clostridia* and related taxa may play a protective role in cardiovascular health.^29–32^ While direct mechanistic evidence remains limited, *Clostridia* are negatively associated with risk factors like obesity, and *Oscillospiraceae* show an inverse relationship with inflammatory states such as IBD.^31^ On genus level, *Eubacterium* has also been inversely linked to atherosclerosis.^30^ Earlier observations linked *ER4 sp900317525* to lower blood pressure variability - a recognized independent risk factor for cardiovascular events, and atherosclerosis.^30–32^ Our results align with these earlier observations.

Earlier studies have suggested that *ER4 sp900317525* may exhibit **broader protective potential** beyond effects on cardiovascular health. *Oscillospira* are considered as potential probiotics.^33^ *Eubacterium rectale* (*ER*) has demonstrated significant systemic benefits, including positive effects on mood and depression.^34,35^ *ER* also appears to play a role in systemic inflammation, with observed reductions in patients with psoriasis and psoriatic arthritis, and protective trends in rheumatoid arthritis.^36,37^ Lower gut abundance of *Eubacterium rectale* was also linked to COVID-19 mortality.^38^ Our findings linking this taxon to reduced all-cause mortality, an effect primarily driven by non-cardiovascular diseases, are in line with these earlier findings and support broader favorable effects of *ER4 sp900317525*.

As to the **potential mechanisms,** SCFAs have been proposed. *Oscillospiraceae* are known butyrate producers and have previously been negatively associated with obesity and inflammatory bowel disease. SCFAs produced by fiber-fermenting bacteria - including members of *Oscillospiraceae* but also *Faecalibacterium*, *Roseburia*, and *Lachnospiraceae* - show anti-inflammatory effects, maintain intestinal barrier integrity, reduce atherosclerosis and contribute to blood pressure regulation, suggesting that specific microbial functions may be cardioprotective.^7,39,40^

### Oral – heart link

*Scardovia* is part of the hypothesized **oral-systemic link,** but previously it has not been specifically linked to CVD.^41^ It can be considered an indicator of general oral dysbiosis, which is a suspected promoter of CVD. One hypothesized mechanism is b**acterial translocation**. Oral bacteria can enter the bloodstream (bacteremia) and contribute to systemic or local inflammation, endothelial dysfunction and arterial plaque formation. As a hardy, acid-tolerant anaerobe, *Scardovia* has the biological resilience required to survive gastric transit and colonize the gut - a profile characteristic of systemic invaders derived from the oral cavity.^25,42^ The role of oral bacteria in MACE is further supported by a parallel study in the Norwegian HUNT cohort (Wolford *et al., submitted*).

*L. gasseri* and other *Lactobacilli* have been detected in saliva samples and described as part of oral microbiota associated with dental caries.^28,43,44^ *Lactobacillus* abundances are increased in the saliva of atherosclerosis patients.^45^ *Lactobacillus* species, including *L. gasseri*, can cause severe infections. *Lactobacillus* has been identified as causative taxa in infective endocarditis, a link supported by both animal models and over a hundred documented human cases^46,47^ The association of *Limosilactobacillus* and *Lactobacillus* with MACE can be considered as a signal of oral ecological imbalance. While these taxa are often viewed through a probiotic lens, their role in the oral cavity appears more complex when linked to cardiovascular risk.

Oral bacteria like *L. gasseri* and *Scardovia* share a common risk factor with MACE: high sugar intake. The observed associations of *Scardovia* and *Limosilactobacillus* with MACE are unlikely to be fully explained by shared cardiometabolic risk factors. Adjustment for metabolic covariates, including BMI, did not attenuate the effects.

Why might microbes normally inhabiting upper GI tract or oral cavity be increased in fecal samples? A plausible explanation for the increased abundances of typically upper gastrointestinal (GI) or oral taxa in fecal samples relates to disruption of normal host– microbiome barriers and ecological balance. One well-established mechanism is **hypochlorhydria**. Reduced gastric acidity, which can be due to proton pump inhibitor (PPI) use or conditions such as chronic atrophic gastritis, weakens the stomach’s bactericidal function. This facilitates survival and downstream passage of acid-sensitive, upper GI–associated taxa (e.g., *Limosilactobacillus* spp. such as *L. antri* and *L. gastricus*), leading to their increased relative abundances in stool samples. An alternative or complimentary explanation is that these taxa act as markers of gut dysbiosis. In this framework, an apparent increase of upper GI or oral microbes in feces reflects depletion of the indigenous colonic microbiota rather than true overgrowth. Perturbations such as antibiotic exposure or intestinal inflammation can reduce the abundance and diversity of resident gut commensals, thereby inflating the relative abundance of transient or non-resident taxa, including *Limosilactobacillus*.

The role of gut dysbiosis and the translocation of oral bacteria as drivers of cardiovascular risk is increasingly recognized.^49,50^ These findings suggest that microbial translocation and the ectopic colonization of the gut by oral or upper-GI inhabitants may exert detrimental systemic effects. Our results contribute and extend the evidence regarding risk, demonstrating that increased fecal abundances of taxa typically associated with the oral cavity, including *Scardovia*, *S. wiggsiae*, and *Limosilactobacillus*, or the upper gastrointestinal tract (*L. gasseri*, *L. antri*, and *L. gastricus*) are associated with a higher risk for MACE and cardiovascular mortality. Conversely, higher levels of *ER4 sp900317525*, a group primarily localized to the intestinal environment, associated with a reduced risk for MACE and mortality.

### Strengths and limitations

The primary strengths of our study lie in its prospective design, large population-based cohorts, and a long follow-up period, which enabled the observation of incident MACE and cardiovascular mortality. These outcomes underscore the clinical relevance of the identified microbial signals, which remained consistent across different taxonomic levels. The findings are further bolstered by deep phenotyping, rigorous adjustment for known classical cardiometabolic and genetic confounding factors and, importantly, successful replication across four independent cohorts in three different countries. The use of shotgun metagenomics in FINRISK study, although shallow, instead of 16S ribosomal RNA sequencing enabled species-level assessment of the microbiota, including less prevalent taxa.

However, several limitations apply. The observational nature of our study precludes causal inference. The assessment of microbiome at a single-time point at baseline does not capture temporal dynamics, whereas the lack of strain-level resolution limits the interpretation of functionally diverse taxa such as *L. gasseri*. Finally, the Nordic-centric population may affect generalizability, and unmeasured confounders could still influence these associations.

## Conclusion

Our study demonstrates that the gut microbiota is significantly associated with the risk of incident MACE. While these findings highlight the microbiome’s potential in risk assessment, clinical trials or animal experiments are necessary to establish causality and determine if microbial modulation can contribute to the prevention of cardiovascular events. Furthermore, as our analysis focused on primary outcomes, the role of microbiome in secondary prevention remains to be assessed. Finally, understanding the biological determinants of microbiome’s effects require further studies assessing microbial functionalities and metabolites, to identify key molecules responsible for the effect.

## Data Availability

Due to GDPR and cohort-specific regulations, individual-level data cannot be made publicly available. Access to FINRISK, SIMPLER, SCAPIS, and HUNT data is available to qualified researchers through the respective cohort data access procedures and required ethical approvals.

## Acknowledgements

The data used for the research were obtained from THL Biobank (study number:THLBB2025_17). We thank all FINRISK 2002 study participants for their generous participation in Finnish health research. We thank the Health and Social Data Permit Authority Findata for providing the health register data (permit number: THL/3273/14.02.00/2025). We wish to acknowledge CSC – IT Center for Science, Finland, for computational resources. We acknowledge SIMPLER for the data access. SIMPLER is funded by the Swedish Research Council (2017-00644 and 2021-00160). The computations and data handling of SIMPLER data were facilitated by resources from the National Academic Infrastructure for Supercomputing in Sweden (NAISS) at the Uppsala Multidisciplinary Center for Advanced Computational Science (UPPMAX) under project simp2025020.

This research has been conducted using the The Swedish CArdioPulmonary bioImage Study (SCAPIS) Resource, under Petition Number 774.

The main funding body of SCAPIS is the Swedish Heart and Lung Foundation. The study is also funded by the Knut and Alice Wallenberg Foundation, the Swedish Research Council, VINNOVA (Sweden’s Innovation agency), the University of Gothenburg and Sahlgrenska University Hospital, Karolinska Institutet and Region Stockholm, Linköping University and University Hospital, Lund University and Skåne University Hospital, Umeå University and University Hospital, Uppsala University and University Hospital. We would like to acknowledge the help of Biobank Sweden and the local biobank facilities for their services in handling of biological samples and biobanking.

The computations and data handling were enabled by resources in project sens2019512 provided by the National Academic Infrastructure for Supercomputing in Sweden (NAISS) at UPPMAX, funded by the Swedish Research Council through grant agreement no. 2022-06725.

